# FAMES: Federated additive model using piecewise exponential survival data

**DOI:** 10.64898/2026.05.15.26353335

**Authors:** Nazmul Islam, Chongliang Luo, Jiayi Tong, Grant Weller, Daniel A. Pollyea, Andrew Kent, Steven Bair

**Author notes:** **Communicating Author:** Nazmul Islam, RefinedScience, 2115 N Scranton Street, Suite 2-70, Aurora Colorado, 80045, USA.

## Abstract

**Introduction:** In analyses of time-to-event data, clinical characteristics can have non-linear impacts on survival outcomes, and understanding this dynamic behavior is crucial for producing real-world evidence (RWE). Nonetheless, estimating these dynamic effects is inherently challenging when utilizing real-world data (RWD), especially since sharing individual-level patient data (IPD) is heavily restricted due to regulatory limitations. Additionally, computational difficulties are exacerbated by the high dimensionality, inter-dependency, rarity, sparsity, and scarcity of features. While data augmentation through collaboration across multiple sites might address these challenges, such collaboration is often infeasible and hindered by regulatory measures that protect patient privacy, thereby preventing the sharing of IPD between sites.

**Objectives:** To address this challenge, we propose a privacy-preserving regularized algorithm that eliminates the necessity of aggregating any protected health information across sites. This algorithm employs a penalized **f**ederated **a**dditive **m**odel utilizing piecewise **e**xponential **s**urvival (FAMES) data and estimates non-linear effects of features while accounting for non-varying confounding effects. The model is flexible and can accommodate both multiple and multivariate smooth effects simultaneously.

**Methods:** The proposed model transforms survival data into a piecewise exponential data (PED) structure and casts the semi-parametric optimization problem into a generalized additive modeling framework assuming Poisson distribution. The model uses orthonormal splines to approximate non-linear effects and incorporates L2-norm based penalty terms to control the smoothness and goodness-of-fit of these effects. The algorithm is optimized using site-specific aggregated summary statistics and is solved iteratively through the Newton-Raphson method.

**Results:** The model is employed to assess the smooth effects of clinical features, such as age and numeric laboratory values, on overall survival using RWD from approximately 874 newly diagnosed Acute Myeloid Leukemia (AML) patients treated at seven distinct sites in the United States. The model exhibited non-linear smooth effects for lactate dehydrogenase, platelets, and others underscoring their strong association with disease prognosis. The model demonstrates a lossless property, providing estimates of smooth and fixed effects that are comparable to those derived from the pooled PED. Additionally, the inference of parameters for testing the nullity of effects remains consistent. This model is communication-efficient, necessitating roughly twelve rounds of communication across sites.

**Conclusion:** We anticipate that this model can facilitate multisite collaboration and enable smaller sites to participate in generating and validating RWE, especially for rare diseases. While the model was applied within the context of AML, it is disease-agnostic and can be implemented in any other clinical context and across various sites globally without losing any generality.

## 1. Introduction

Cox proportional hazard (Cox-PH) regressions with varying coefficients quantify non-linear effects of clinical features on survival endpoints. These models are preferred over linear models for understanding dynamic features’ behavior in generating real-world evidence (RWE) from real-world data (RWD), especially for rare blood malignancies like acute myeloid leukemia (AML), where biological processes and disease progression change dynamically and rapidly. While these models are flexible, the computational challenges in estimating model parameters may stem from high dimensionality, inter-dependency among features, sparsity of covariates, rarity of events, data scarcity, and other pertinent RWD issues. Although data augmentation through multi-site collaboration may alleviate these estimation issues at an extent, such collaboration is often impeded by regulatory measures protecting patient privacy, which obstruct sharing individual-level patient data (IPD) across sites.

To address the challenges associated with data sharing, federated models have been investigated and implemented in recent studies for various prediction, causal, and association problems across diverse data modalities and response distributions^1-4^. However, the application of federated modeling for time-to-event outcomes, particularly in the context of RWD for rare diseases, has not been thoroughly explored. Different multivariable federated Cox-PH models have been developed focusing on the estimation and inference of non-varying fixed effects of features^5-10^. In particular, federated regressions for horizontally distributed survival data have been developed through surrogate likelihoods by utilizing aggregated gradients and Hessians from each site or alternatively by transforming time-to-event data into a classification problem^11-13^. Federated survival modeling upon transforming data into a Poisson modeling framework has also been explored and non-varying effects of covariates are estimated by optimizing the corresponding likelihood function^10,12^. Very recently, federated modeling techniques for fitting additive models adjusting for varying effects of covariates under generalized modeling framework for non-survival data have been studied. In a similar vein, federated generalized additive model^14^ (GAM) for location, scale, and shape (GAMLSS) has been proposed to estimate age-specific percentile curves for clinical parameters^15,16^. However, these models are not appropriate for survival data and are not intended for estimating multiple or multivariate smooth effects while accounting for the effects of non-varying confounders, which is the primary focus of this paper.

As the dimensionality of available clinical data increases, models that account for the aforementioned complexities are increasingly important to guide clinical predictions and decision making. For an instance, varying effects may signal how the association of a numeric feature (e.g., age, blasts) relates to survival and whether the corresponding effect remains constant or changes across the underlying covariate range or time. Allowing these effects to vary smoothly over one or more dimensions can also uncover clinically meaningful thresholds, non-linearities, and interaction patterns that would be obscured under standard linear or categorical modeling with fixed effects. These smooth, potentially multidimensional effects can also assist in selecting optimal cut-off values for laboratory measurements (e.g., platelet counts) that are clinically significant and can be used to refine clinical response definitions based on these lab values. Furthermore, such dynamic estimates increase the granularity with which a clinician can predict the impact of a specific patient’s pattern of clinical data parameters, and the influence on outcomes of a specific data parameter when varying from patient to patient, ultimately helping pair the right patient with the right treatment modality. While such dynamic models are powerful to generate insights, these typically require a larger heterogeneous dataset and collating such dataset by pooling IPD across sites under rare diseases is often challenging. When pooling IPD is not feasible, a two-stage IPD meta-analysis can be adopted to estimate varying effects. However, this method may produce sub-optimal results, especially when the number of sites is limited and each site has a relatively small patient population with varying distribution in covariate space. In the federated model literature, privacy-preserving models that allow for the estimation and inference of smooth effects are limited and, to the best of our knowledge, have not been explored in detail. To mitigate this gap, we propose a privacy-preserving regularized algorithm that obviates the need to pool any IPD by fitting a penalized **f**ederated **a**dditive **m**odel exploiting piecewise **e**xponential **s**urvival (FAMES) data and allows to estimate non-linear effects of features while controlling for non-varying confounding effects. The proposed model casts survival regression within the GAM framework by converting time-to-event data into a piecewise-exponential data (PED) structure while approximating smooth effects via common basis function expansion across sites. We applied two sets of transformation to induce orthogonalization of bases and diagonalization of penalty terms. Due to the separability of the underlying log-likelihood across independent sites, the global gradient (the first derivative of log-likelihood) and Hessian (the second derivative of log-likelihood) are algebraically equivalent to the sum of site-level gradients and Hessians. This allows for precise federated Newton-Raphson updates without the need to share IPD leading to numerically identical results to the pooled analysis. The novelties of the proposed method include: (a) the simultaneous estimation of fixed and smooth effects using only summary information (non-IPD) from local sites, (b) a modeling mechanism allowing to account for multiple smooth effects simultaneously, (c) a flexible modeling framework that allows for interaction of smooth effects with more than two dimensions, and (d) the quantification of uncertainty for both fixed and varying effects.

The proposed model is disease-agnostic and applicable to any horizontally distributed survival data. The numerical performance of FAMES was evaluated using RWD from 874 AML patients treated with de-novo venetoclax and azacitidine (ven/aza) at seven geographically dispersed sites across the United States. Parameters were estimated using both pooled and federated approaches, and the results were compared.

## 2. Methods

### 2.1 Description of datasets

The primary site, designated as the orchestrator, consists of 359 adult patients newly diagnosed with AML who received treatment with ven/aza at the University of Colorado (CU) Cancer Center between 2015 and 2024. Data from 32 secondary sites were sourced from the Flatiron Health dataset, a licensed, de-identified dataset containing patient information for individuals diagnosed with AML using ICD-9 and ICD-10 codes. These individuals had a minimum of two documented clinical visits between 2014 and 2023 and received ven/aza as frontline therapy. During the study period, the de-identified data were collected from approximately 280 cancer clinics across the United States, encompassing around 800 sites of care, primarily within community oncology settings. The data were de-identified and subject to obligations to prevent re-identification and protect confidentiality. Patient-level baseline data (prior to treatment start date) were discussed previously^10,17,18^ and thus the details are omitted here; see Supplemental Figure 1 for summary statistics of features. For our application, we excluded sites with fewer than 50 patients, resulting in a final number of seven participating sites, including the orchestrator having 874 AML patients. The datasets are horizontally distributed, meaning the same set of features is available across all sites. The analytical dataset includes demographic information (e.g., age, sex), laboratory data (e.g., white blood cell (WBC), absolute neutrophil count (ANC), platelets (Plt), hemoglobin (Hgb), blasts, lactate dehydrogenase (LDH), and prior history of medical comorbidities (e.g., chronic kidney disease (CKD), hypothyroidism, hypertension, heart disease, coagulopathy, non-AML cancer, myelodysplastic syndrome (MDS), chronic obstructive pulmonary disease (COPD), venous thromboembolism (VTE), and obesity). Additionally, two patient risk classification models, the refined European Leukemia Net-2024 (rELN24)^19^ and the refined risk model (RRM)^18^, were used as potential confounders to characterize the effects of genetic features on survival. Missing laboratory values and genetic features were imputed locally using the median of the corresponding lab values and mode of categorical features, respectively. Unlike the data in CU, patient-level blasts were recorded as a categorical value with grid of intervals (e.g., 0-5, 6-10, … 50-100, etc.) in Flatiron. To consolidate, these percentages were transformed into a numerical number by taking a random sample from a uniform distribution of the minimum and maximum values of corresponding intervals for each patient. It is important to note that although accessing to these Flatiron secondary sites’ datasets is available, the datasets were treated as siloed and locked, ensuring no sharing of IPD across sites.

### 2.2 Additive model for survival data

Let *δ*_*i*_, ***X***_i_ and ***Z***_*i*_ be the event indicator, *P*-dimensional non-varying covariate vector, and *Q*-dimensional varying covariates for the *i*-th subject, respectively. Define *κ*_*i*_ be the time-to-event for the *i*-th subject. Similarly, censoring time is denoted by *c*_*i*_. Denote the survival data where *t*_*i*_ = *min*(*κ*_*i*_, *c*_*i*_). The hazard function under the Cox-PH is written as

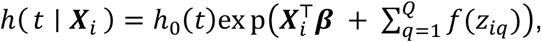

where *h*_0_(. ) is cction and ***β*** is a set of unknown parameters quantifying the association between features and survival end-points and *f*(. )’s refers to smooth effects of ***Z***. We leverage the statistical equivalence between the Cox-PH model and the piecewise exponential additive model (PAM) assuming *h*_0_(. ) is approximated as a constant over fine-grid of time intervals^20^. For the convenience of illustration, we assume *Q* = 1. We approximate the smooth effect *f*(. ) by splines as

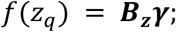

where ***B***_***z***_ represents the B-splines or truncated power splines (TPS) basis functions and ***γ*** represent the corresponding (*l* × 1) vector of unknown basis coefficients.

We cast the model into a semi-PAM framework utilizing the PED structure with design matrix defined by ***X***_*i*_ and ***Z***_*i*_. Let the PAM define by

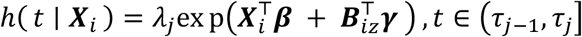

where *λ*_*j*_ is constant within each discrete time intervals. Note that when intervals (*τ*_*j*−1_, *τ*_*j*_]’s, *j* = 1, …, *J*, are short and dense enough, this model approaches the Cox-PH model and yields nearly similar parameter estimates^21^. We fit the model via GAM framework by maximizing the Poisson likelihood function assuming log link. Let the number of events for the *i*-th subject at the *j*-th interval be

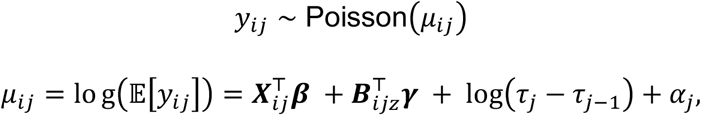

where *log*(. ) is an offset term which is the interval length serves to account for time duration each subject is at risk in each interval and *α*_*j*_ = *log* ( *λ*_*j*_ ) is log baseline hazard for the *j*-th interval.

### 2.3 Federated penalized piecewise exponential additive model

When sharing IPD is challenging, fitting a global survival model becomes difficult. Alternatively, one can fit a PEM with Poisson link function using an L2-norm penalty and estimate unknown coefficients by maximizing the corresponding penalized likelihood function. In federated settings, each site’s proprietary data remains secure and locked, so sites only share local summary statistics or sufficient statistics with the orchestrator. Subsequently, these are used to generate a global likelihood function, gradients, and Hessians through aggregation, thanks to the *separability* property. Figure S1 below highlights the steps of fitting PAM in a distributed setting. For implementation, we use truncated linear splines (TLS) to approximate smooth effects. Let ***n***_*k*_ be the number of subjects in the *k*-th site where *k* = 1, …, *K*, ***X***_*k*_ be the (***n***_*k*_ × *P*) design matrix of covariates with row being a (*P* × 1) vector of ***X***_*ik*_′*s*, ***B***_*k*_ be the (***n***_*k*_ × *l*) design matrix of raw basis functions corresponding to the smooth effect, ***y***_*k*_ be the ***n***_*k*_ × *J* vector of *y*_*ijk*_′*s*, and ***o***_*k*_ is the offset term. We assume independence between ***X***_*k*_ and ***B***_*k*_. To adjust for collinearity among features and to control the smoothness of varying effects, L2-norm based block diagonal penalty matrices were used resulting in a penalized Poisson GAM^22^. The corresponding penalized likelihood function can be expressed as

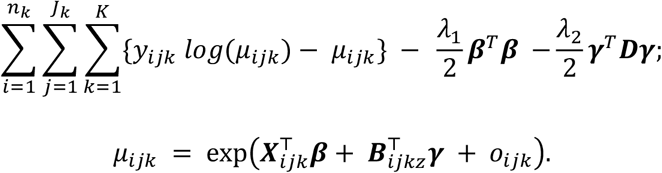

where *λ*_1_ and *λ*_2_ correspond to the ridge and smoothing penalty, respectively. Here ***D*** corresponds to the penalty matrix with first or second order finite-differences applied to spline coefficients; alternatively, second derivatives of the basis function *f*(. ) integrated over the domain of spline can also be used. In our applications, we used first-order difference penalty to define ***D*** for the simplicity of exposition.

### 2.4 Approximation using orthonormal basis functions

Borrowing the idea of orthonormalization, we transformed the basis vectors into orthonormal bases so that they are mutually orthogonal and each has unit length^23^. Orthogonalization has numerical advantages over using just truncated power series since numerical errors, collinearity among TPF, and data imbalances may contaminate both gradient and hessian leading to significant numerical challenges specially in federated model where data are not pooled.

After aggregating all sites’ basis cross-products, a global matrix of basis function (Gram matrix) is computed. The orchestrator applies eigen decomposition on the global cross-product as

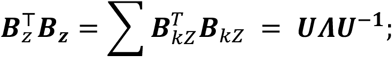

where ***U*** corresponds to eigenvectors (signaling directionality in space) and ***Λ*** to eigenvalues (weights of the corresponding eigenvectors). Note that eigen components with numerically negligible eigenvalues (e.g., <1e-8) are excluded to remove non-singular directions from the spline space. To transform the global basis orthonormal, each site’s basis function is multiplied by ***A***_***z***_ = ***UΛ***^**−1/2**^ such that

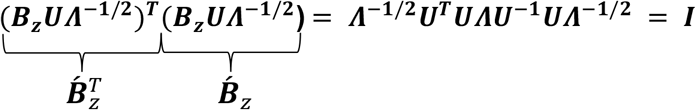

where the transformed site-specific basis functions will be 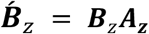 ignoring the superscript *k*. This leads to 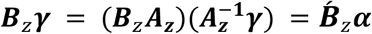 where ***α*** is a vector of coefficients corresponding to orthonormal basis. Subsequently, the penalty term associated with the smooth effect can be re-parametrized as

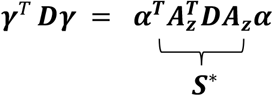

Therefore, the new design matrix becomes 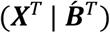 suppressing the subscripts for notational brevity. Next, we can maximize the re-parametrized penalized likelihood function to estimate ***β*** and ***α***.

### 2.5 Construction of Demmler–Reinsch basis functions

The non-diagonal penalty structure ***S***^**∗**^ presents numerical challenges and implementation difficulties during federation, particularly when inverting the high-dimensional Hessian matrix in the Newton-Raphson steps. To resolve, we transform the orthonormal basis into Demmler-Reinsch (DR) basis^24^ so that the penalty structure becomes diagonal. This extra step will induce more numerical stability, simplicity in implementation, and communication efficiency. In this pursuit, an eigen decomposition of ***S***^**∗**^ = ***VΣ***_***s***_***V***^***T***^ is performed where ***V*** is orthogonal (***V***^***T***^***V*** = ***I***) and ***Σ***_***s***_ corresponding to a (***l***′ × ***l***′) diagonal matrix with positive eigenvalues. Directions associated with very small eigenvalues (<10^-8^) signaling noise are dropped. It follows that the penalty term can be re-expressed as below-

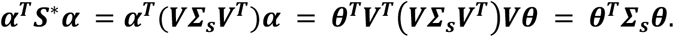

Here ***α*** = ***Vθ***. This leads to DR basis as 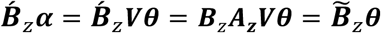 where 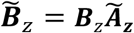 and 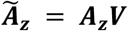. It can be further shown that this reconstructed basis is orthonormal (i.e., 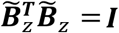). The new design matrix takes the form of 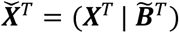 which we use to optimize the penalized likelihood function in estimating the set of parameters 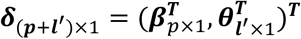 where ***l***′ ≤ ***l***. Note that smooth splines are represented in the DR-basis with a full-rank ridge penalty applied to all corresponding basis coefficients, ensuring that there is no unpenalized null space; this approach guarantees the identifiability of smooth effect estimation without imposing additional constraints in the optimization^25^.

### 2.6 Estimation of federated penalized additive model

The proposed estimation procedures of FAMES are illustratively depicted in Figure 1. It is imperative for the orchestrator and each collaborating site to establish a clear objective and a written protocol that delineates the steps for standardization, normalization, and definition. Any deviations from the protocol, whether in analytical plans or in the definitions of features, must be consolidated or at least acknowledged in advance to ensure valid interpretation of results. It is crucial to employ the same global knots and basis across all sites, broadcast the same global DR-transformation matrix necessary for orthogonalizing the basis to all local sites, and apply the same penalty terms globally to maintain separability. Additionally, observations across sites must be independent. The orchestrator shares the parameter space as a fine grid of potential values. Each site provides summary statistics, including local gradients, Hessians, and local likelihood values. The orchestrator aggregates local information to generate global gradients, Hessians, and likelihood values, which are iteratively used to optimize parameter estimation for each combination of tuning parameters. Multiple communication rounds may be warranted to obtain parameter estimates for each tuning parameter combination. It is important to note that these communication rounds are asynchronous and independent, meaning the sequence of information flow does not affect the optimization process, and no inter-site communication is required. To minimize communication rounds and select tuning parameters, sites perform and update local summary statistics for each tuning parameter combination and share them simultaneously in a sequence with proper labeling, enabling the orchestrator to track them efficiently. The orchestrator performs optimization for each of these values separately but concurrently. Once parameter optimization rounds are completed for each tuning parameter, the orchestrator selects the optimum tuning parameter using the Akaike Information Criterion (AIC) or Bayesian Information Criterion (BIC), which can be derived by aggregating the summary information from local sites. A more detailed explanation of the steps is provided in Table 1.

**Figure 1.**
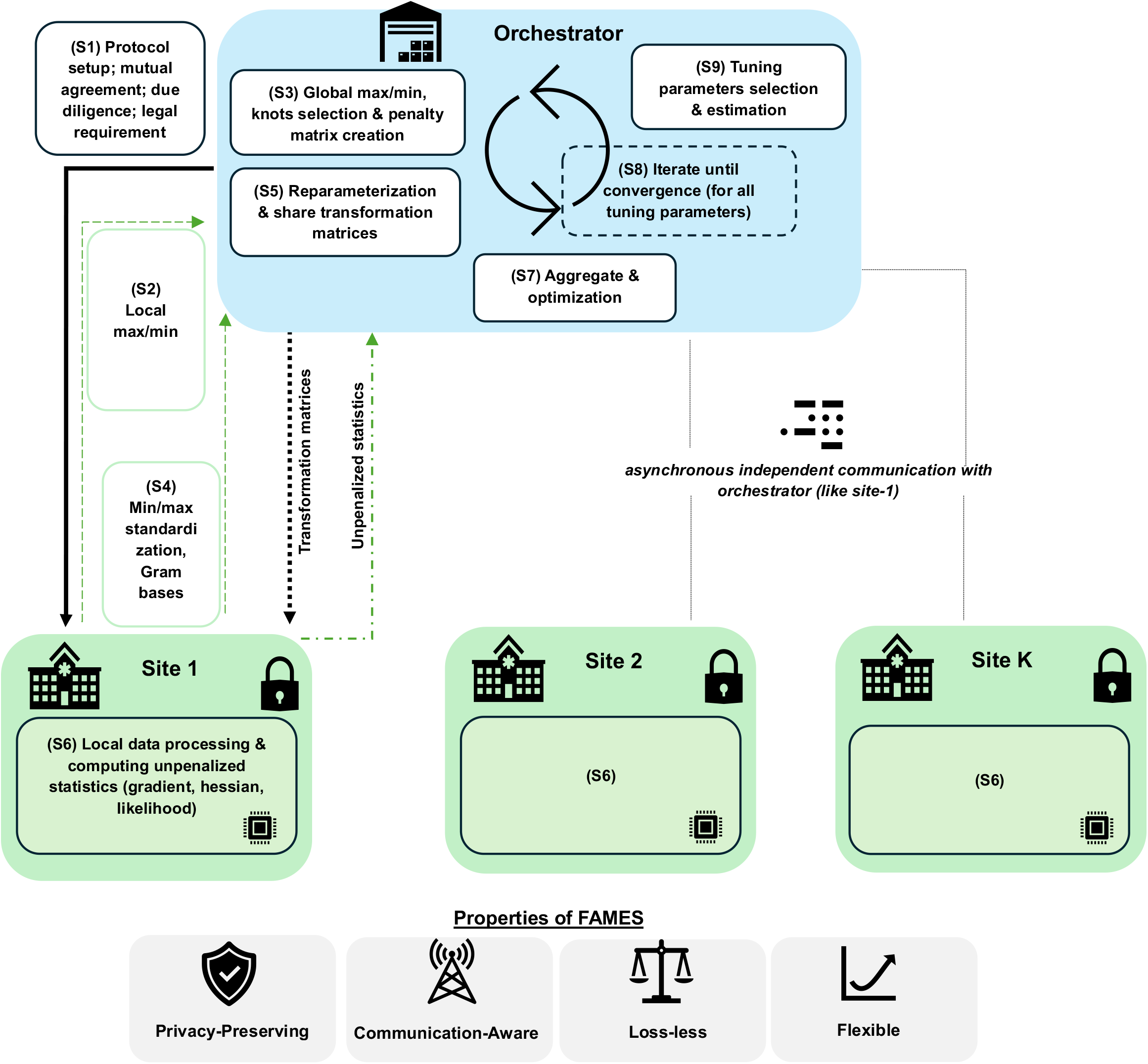
A schematic diagram of FAMES algorithm.

**Table 1.**
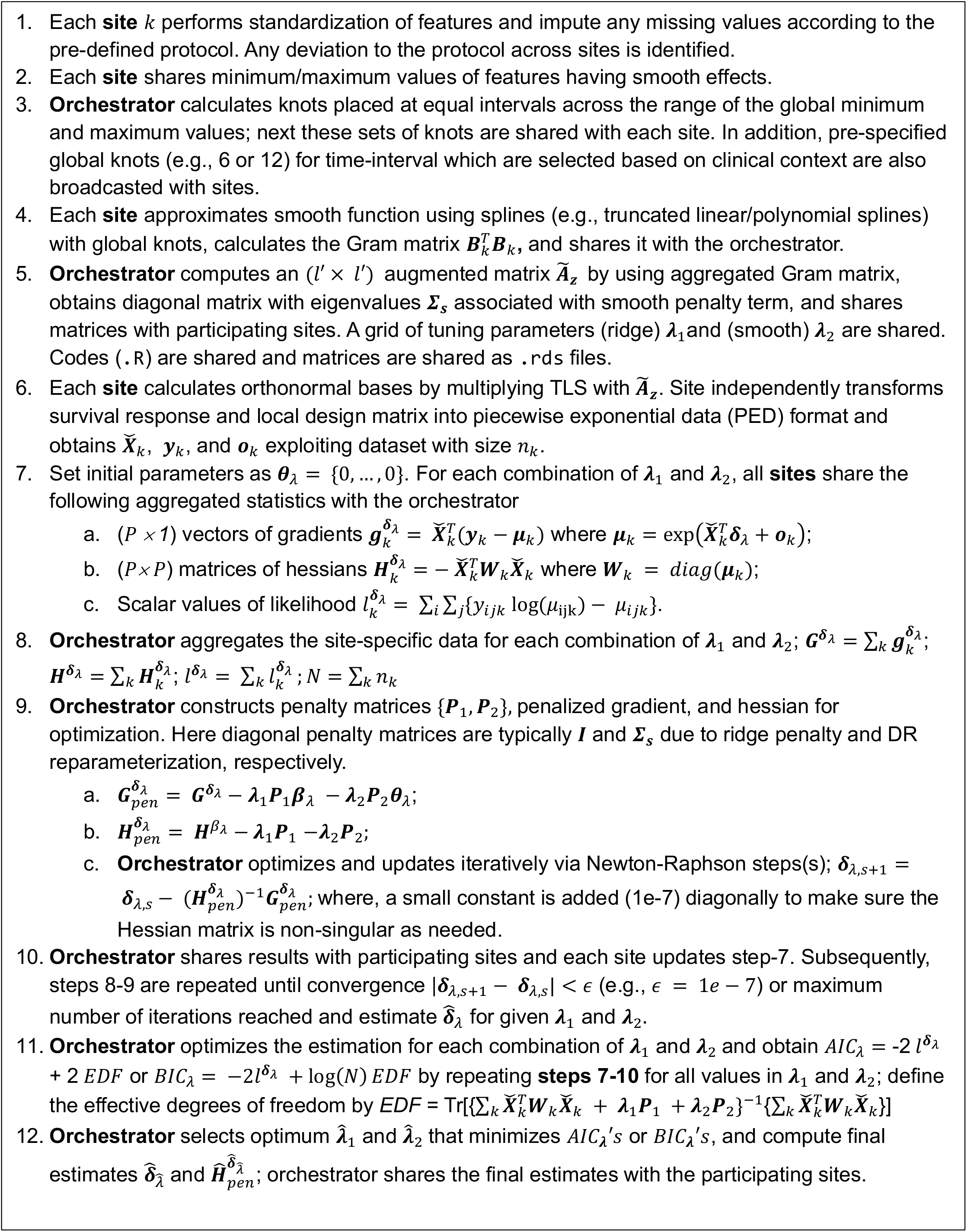
An overview of the FAMES estimation process.

### 2.7 Uncertainty quantification of effects

Fisher information matrix was calculated, and the corresponding standard errors of 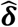 were derived using Moore-Penrose G-inverse of 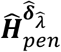 to avoid numerical issues related to singularity. Final smooth effects are estimated as 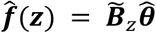 where, 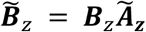 and 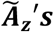 are shared by the orchestrator to each participating sites. The corresponding variance-covariance matrix for each smooth effect is computed as 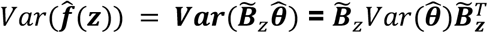. Subsequently, 80% and 95% confidence intervals were computed using Wald-type formula.

### 2.8 Ethical considerations

This retrospective study was approved by the Colorado Multiple Institutional Review Board (COMIRB), study number 23-2059. All methods were performed in accordance with the relevant guidelines and regulations, including the Declaration of Helsinki. Due to the retrospective nature of the study, COMIRB waived the need of obtaining informed consent.

## 3. Results

In our numerical experiment, the primary aim was to estimate the smooth effects of age, WBC, LDH, Plt, Hgb, blast, and ANC, while adjusting for potential confounders such as risk stratification models (rELN24 and RRM), comorbidities, and demographic characteristics on overall survival (OS). In a horizontal setting, features remain consistent across seven sites, including the orchestrator. We have access to the pooled sites, and results are compared against the pooled analysis concerning coefficient estimates and standard errors. The results were derived from seven sites, each comprising at least 50 patients; details of the patient cohort were described elsewhere^10^. Figure 2 summarizes the distribution of numerical features for which smooth effects are warranted. To ensure a fair comparison between competing models, the pooled model was fitted as a PAM with a Poisson link function and implemented as a GAM model using the same penalty structure as employed in the FAMES by specifying parametric penalty terms via `paraPen` in the mgcv^26^ R-package.

**Figure 2.**
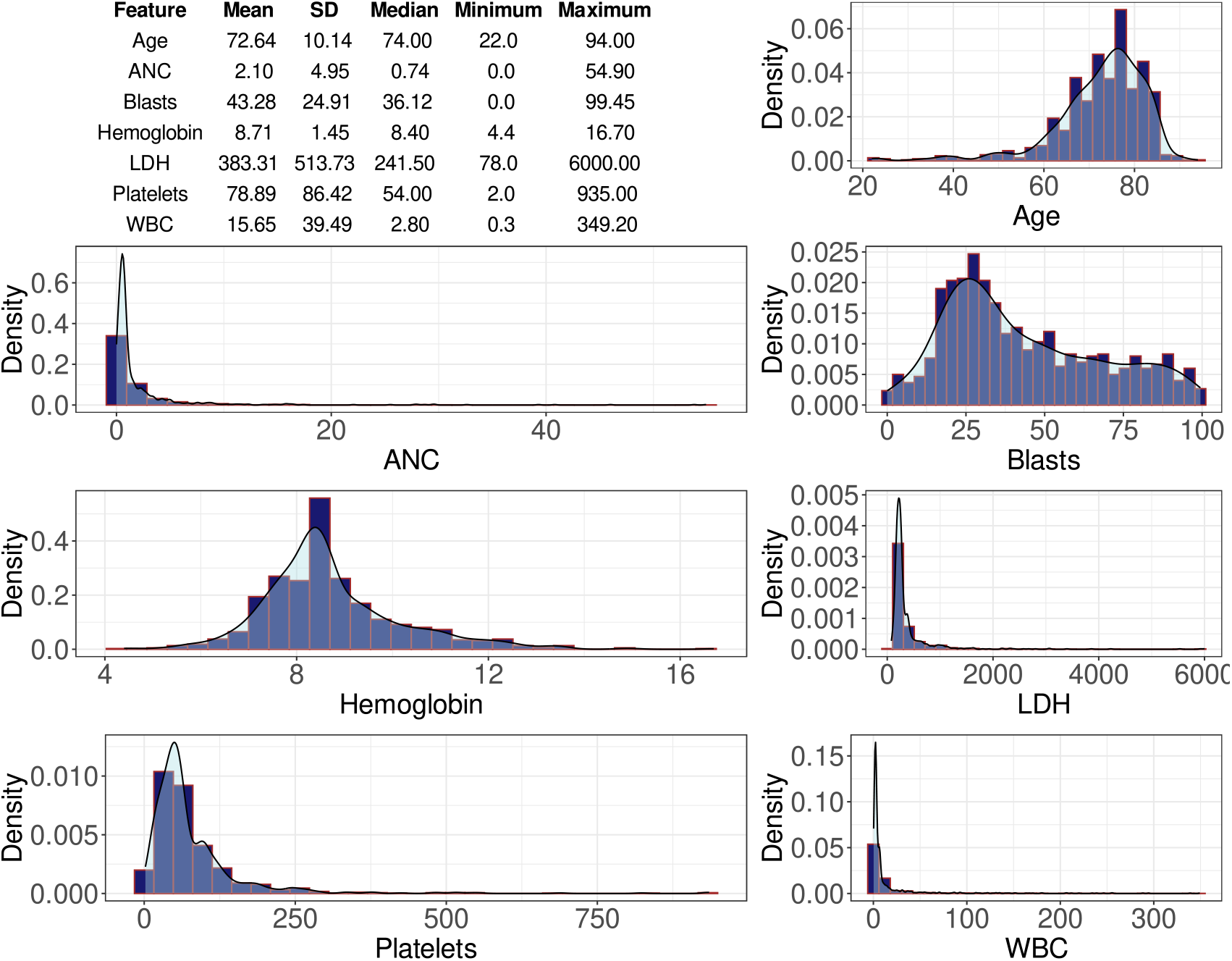
Distribution of numeric features. Reported are the results corresponding to age, ANC (y), absolute neutrophil counts (10^9^/L), blasts (%), hemoglobin (g/dL), lactate dehydrogenase (U/L), platelets (10^9^/L), and white blood cell (10^9^/L).

Figure 3 illustrates a comparative analysis of the outcomes derived from federated and pooled methodologies; reported are the log hazard ratio of covariates and basis coefficients. The federated estimator is algebraically equivalent to the pooled maximum penalized likelihood estimator, resulting in identical point estimates and standard errors. This equivalence underscores the lossless characteristic of FAMES. Optimization was performed simultaneously for all combination of tuning parameters; on average, approximately 12 communication rounds are needed to attain optimal estimates for a given tuning parameter. However, the number of communication rounds is contingent upon parameters such as the number of knots in smooth effects, the number of knots of time intervals, the number of confounders, and the convergence criteria. Various settings were tested, and the results were numerically very similar, although the computation time varied significantly.

**Figure 3.**
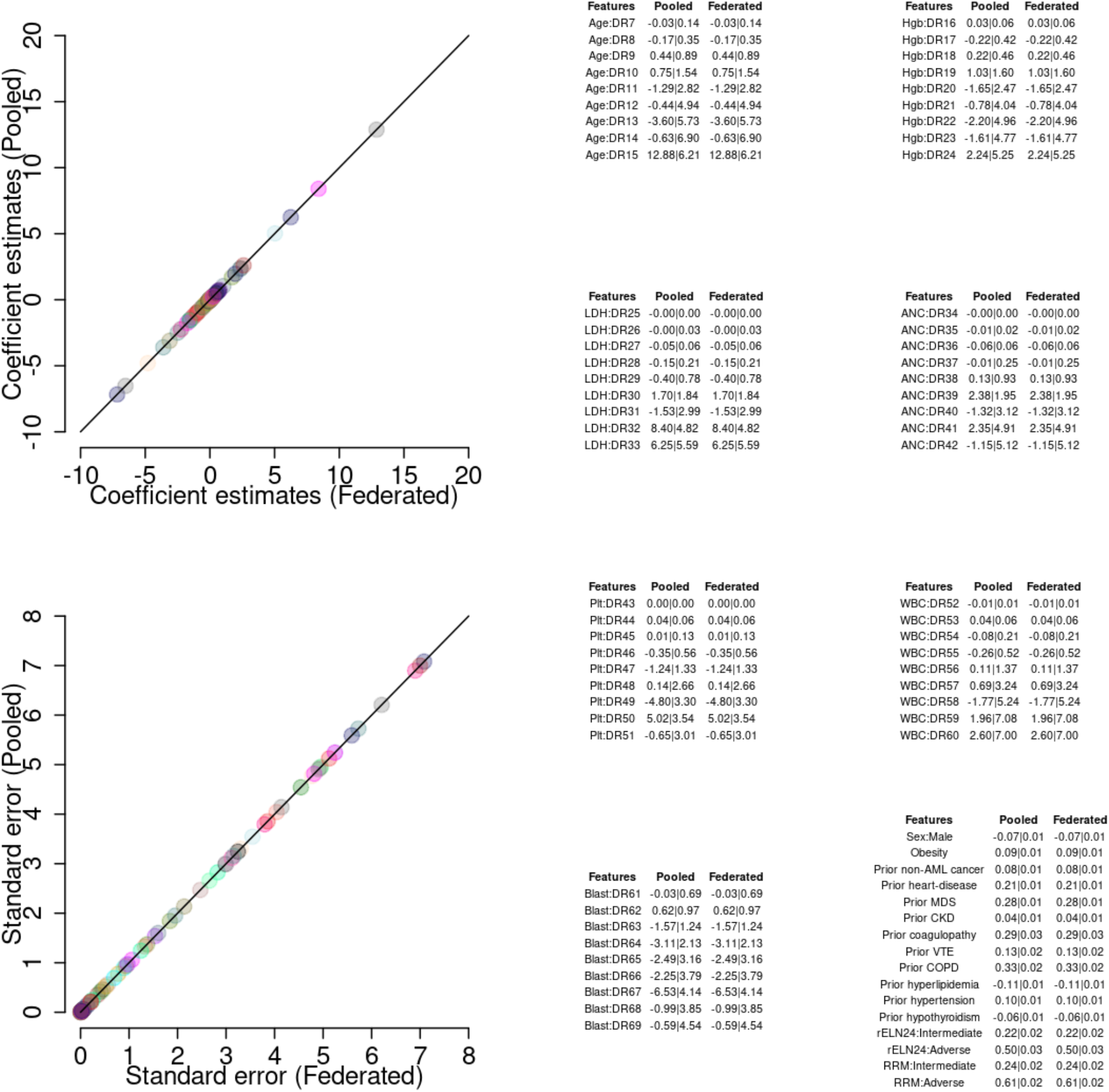
Comparison between federated and pooled analyses. Reported are the coefficient estimates (top-left) along with standard errors (bottom-left). In the right, DR-basis coefficient estimates are provided in tables based on federated and pooled method.

Figure 4 illustrates the smooth effect estimates accompanied by 80% and 95% CIs. The majority of these smooth effects suggest non-linear relationship with survival status. For example, the impact of aging on AML survival appears to progress linearly across a range of age values, with patients over 75-year-old being significantly at risk. Both low and high platelet values demonstrate a negative correlation with OS, whereas the smooth effect of LDH follows an inverted U-shape, indicating that elevated LDH values are adversely associated with outcomes. The effect of baseline blasts exhibits dynamic changes; smaller values seem to correlate positively with OS, while larger values show an inverse association with survival. Nevertheless, a smaller subgroup within the middle range shows a positive correlation with OS. However, it is important to recognize that laboratory value distributions differ across ranges, which may result in fewer patients in extreme ranges, likely leading to overestimation; see Figure 2. Such uncertainty is encapsulated within the confidence intervals and therefore the point estimates need to be interpreted with caution.

**Figure 4.**
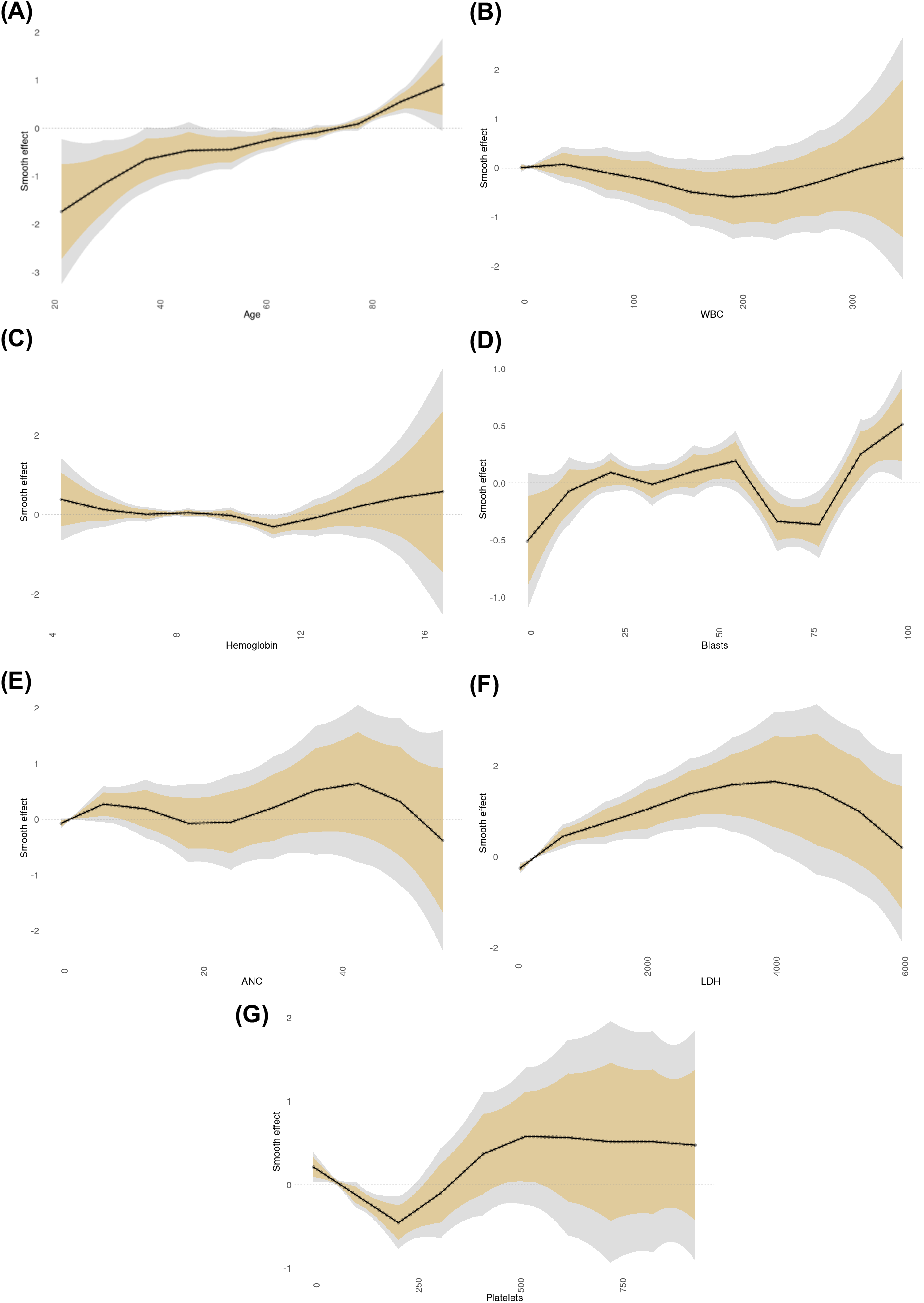
Varying coefficient estimates based on FAMES. Reported are the smooth effects (log hazard ratio) along with 80% (golden) and 95% (gray) confidence intervals for 100 unique lab values that are uniformly distributed within range of the corresponding numeric features for age (y), (B) white blood cell (10^9^/L), (C) hemoglobin (g/dL), (D) blasts (%), (E) absolute neutrophil counts (10^9^/L), (F) lactate dehydrogenase (U/L), and (G) platelets (10^9^/L).

Figure 5A-B demonstrates the combined impact of WBC and LDH on OS using a bivariate smooth function, as outlined in the model described in Supplementary Table 1. This two-dimensional surface estimates how these variables jointly contribute to the log hazard, while keeping the effect of other covariates constant. The highlighted rectangle indicates the common area where the distribution of features is more prevalent, resulting in a tighter CI for the estimated surface in this region (Figure 5C-D). Figure 5E depicts the width of the CI, highlighting the uncertainty levels across the distribution of lab values, which further emphasizes the robustness of the estimates. The smooth function is entirely bivariate, permitting the effect of LDH on hazard to vary with WBC levels, and vice versa. For instance, at a WBC value of 90 (10^9^/L), the effect of LDH values ranging from 245 (U/L) to 1800 (U/L) demonstrates varying effects on OS, indicating a transition from favorable to unfavorable associations as value increases. In a similar manner, the surface plot can be interpreted for any other values in either direction, indicating that the Log HR dynamically varies across the feature space. A separate model was fitted to estimate the combined effect of age and blasts, and the corresponding estimates of the two-dimensional surface are illustrated in Supplementary Figure 2.

**Figure 5.**
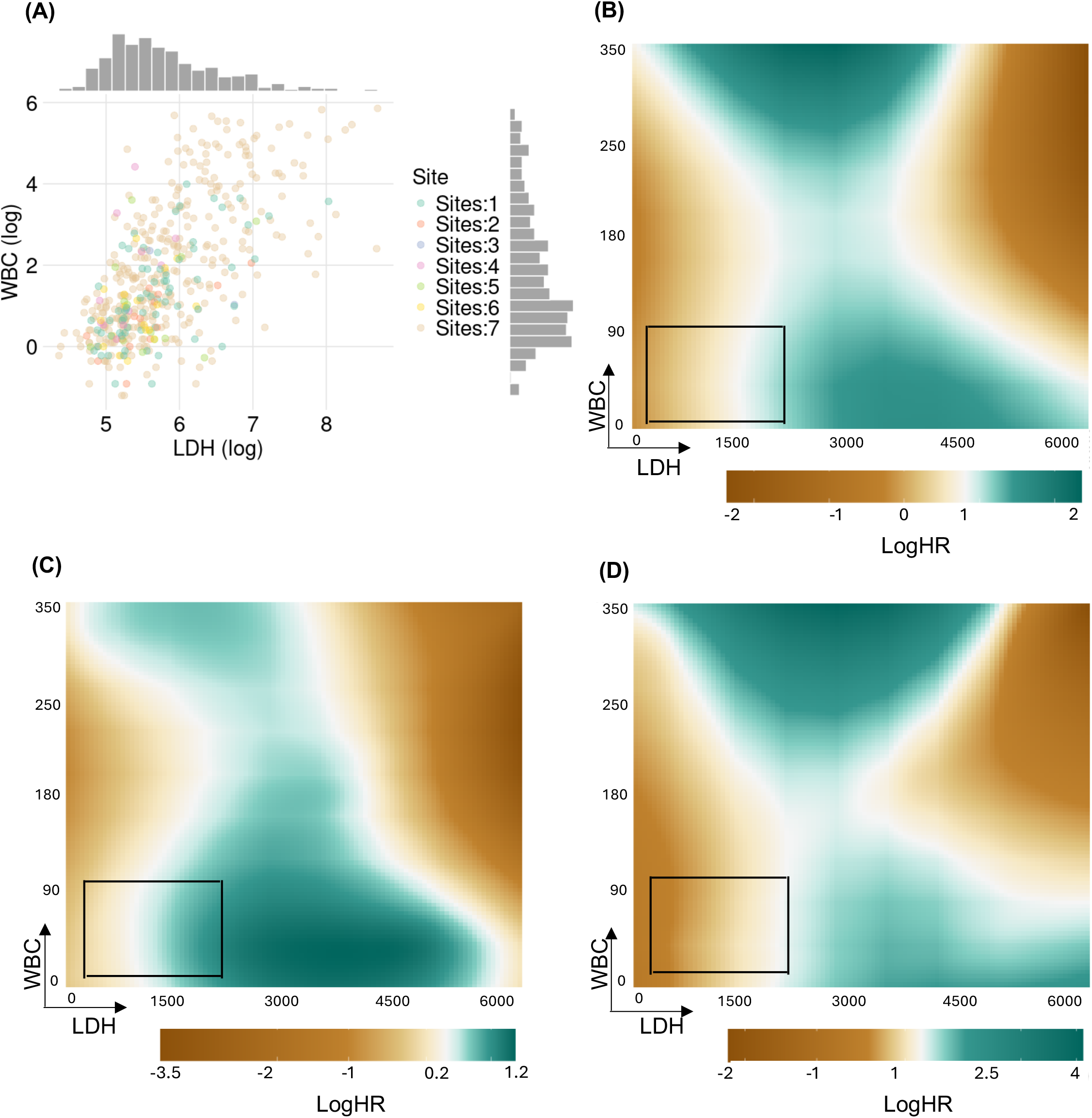
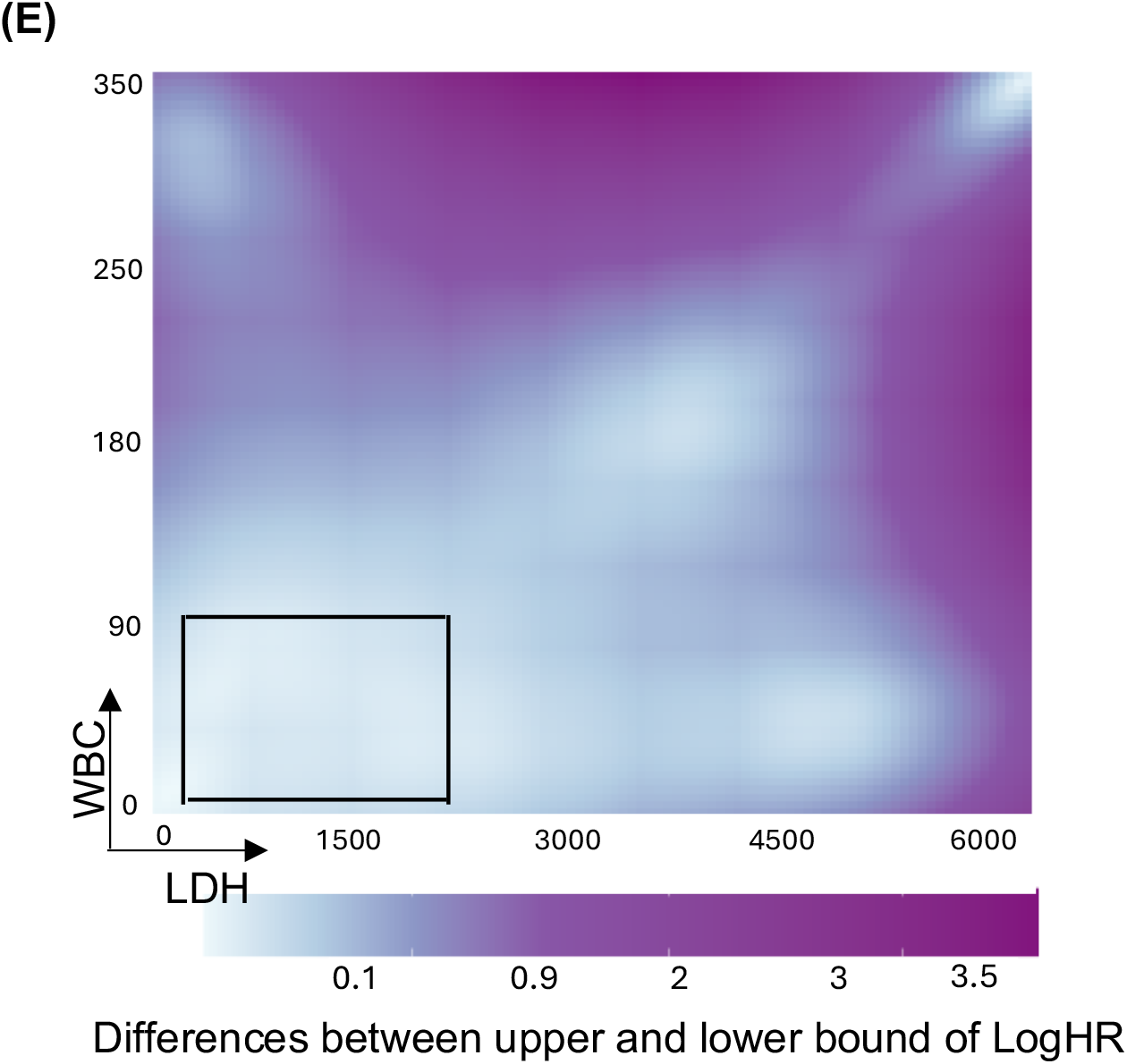
Estimates of bivariate smooth effects. Reported are the interaction effects (log hazard ratio) between WBC and LDH that are uniformly distributed within the minimum and maximum values of the corresponding features. Heatmaps correspond to (A) distribution of WBC and LDH values across 7 sites, (B) point-estimates, (C) lower-bound and (D) upper-bound of 95% CI, and (E) Width of confidence intervals. The highlighted box corresponds to the area with high concentration of data for LDH = {240, 1800} and WBC = {1, 90}.

## 4. Discussion

In our previous work, we have developed a federated algorithm called FedPPEM, for Cox-PH model with no specification of the non-linear effect terms. In this paper we present a federated additive algorithm FAMES to estimate non-linear effects by including flexible smoothing spline terms in the models. Our results of the estimated smooth effects of age and six clinically important lab signal non-linear effects of these dynamic variables in studying the progression of AML. Being able to pinpoint where a patient lies across multiple clinical variables and better predicting how together they will influence response to therapy or overall treatment outcomes, empowers the clinician to better care for each patient. Another key contribution of this study is empirically demonstrating that the federated estimator is algebraically equivalent to the pooled maximum penalized likelihood estimator under certain conditions, such as identical basis expansion, knots, and design matrix construction. Because the Poisson log-likelihood is separable across independent observations at different sites, the global gradient and Hessian are exactly the sums of their site-level counterparts, leading to *loss-less* estimation properties. Given that this summary statistics are sufficient for estimation and inference, the proposed framework is not only statistically efficient but also preserves *patient-privacy*, as no IPD is needed to execute this algorithm.

In a federated setting, estimating smooth effects poses numerical challenges, especially when aggregating Gram matrices from diverse sites. To tackle these challenges, a two-step transformation process was employed: first, the spline basis was orthonormalized, followed by a rotation to align coordinate axes with the penalty eigendirections obtained through the eigen-decomposition of each penalty term. Near-zero eigenvalues were discarded before applying the ridge penalty, thus eliminating near-singular directions from the spline space and ensuring that the penalty is full-rank and diagonal in the DR basis. This approach guarantees the strict convexity of the penalized objective and a unique solution without requiring additional centering or identifiability constraints on the smooth terms. Two optimizations were conducted: initially, for each combination of tuning parameters, coefficient estimates were optimized, and this process was repeated for all combinations; subsequently, the final estimates corresponded to the optimal tuning parameter were selected based on AIC/BIC values. The models were designed to be *communication-aware*, necessitating an average of 12 rounds of asynchronous communications between sites and the orchestrator. Furthermore, the model exhibits *flexibility* by accommodating various functional forms of smooth effects, and it facilitates the adjustment for multiple smooth effects as well as multidimensional regression surfaces.

While the model demonstrates promising results, it does have certain limitations and potential areas for future research that could enhance the modeling framework. A) The model presumes that the PED with Poisson regression can effectively approximate the Cox-PH model, which may be a strong assumption in some instances and could be violated in heterogeneous RWD. B) As with any federated model, achieving absolute accuracy in standardizing features across sites is challenging. For implementation, it may sometimes be necessary to relax data definitions; however, this strongly depends on the objective, significance of the problems, impacts of results, and the underlying context. C) Additionally, enumerating patient overlap across sites is difficult. However, in our application, data were non-overlapping. We were able to validate this as we had access to pooled datasets from all seven sites. D) The model may not be optimal for sites with a limited number of patients, as such small sample sizes can result in sparse or poorly conditioned local Hessians. Furthermore, extrapolating and interpolating to generate a smooth curve may be suboptimal in regions where the distribution of features is less prevalent. These areas can typically be captured through standard errors; therefore, smooth curve point estimates should be interpreted with caution in conjunction with CIs. E) The proposed model does not explicitly consider heterogeneity across sites, which may require incorporating site-specific coefficients in the FAMES model to account for site-specific variability in response and attributes. F) The model is not designed for causal modeling or prediction problems, which we defer for future research. G) The model assumes independent observations across sites; thus, the proposed algorithm cannot be applied in a longitudinal or functional setting where covariates are observed stochastically at fine-grid of intervals or time points for each patient. Such settings require appropriate enumeration of Hessians to account for inter-subject dependency, which, if not properly addressed, may lead to erroneous inference. Therefore, another significant future direction of our research involves extending FAMES to accommodate longitudinal and functional data.

## 5. Conclusion

The proposed FAMES facilitates collaboration across multiple sites in the pursuit of generating clinical insights while ensuring the protection and privacy of proprietary data within each institution’s system. The model offers a practical solution for estimating the dynamic effects of biomarkers across sites without sharing IPD. By incorporating a diverse patient population from various locations, this approach enhances inclusivity and results in a more reliable and robust estimation of population-level parameters. The model can significantly benefit the estimation of dynamic effects of biomarkers for under-represented subgroups.

## Acknowledgments

The authors express their sincere gratitude to Justin Dale, Jamie Reuben, James Coates, Alex Williams, and Karan Sapiah from RefinedScience for their invaluable assistance in processing, extracting, and organizing the clinical datasets from the University of Colorado. Additionally, we extend our thanks to Ujjwal V. Kulkarni and Frank Markson from RefinedScience for their efforts in establishing the computing environment.

## Author contributions

NI and CL designed the study and drafted the manuscript. NI processed the structured datasets for analysis and performed numerical analyses. JT, GW, DAP, AK, and SB edited the draft and interpreted the results of the analyses. All authors reviewed, provided constructive comments, and agreed to its publication.

## Supplementary material

Supplementary material is available along with the submission with additional methods and numerical results pertinent to the study.

## Data availability

The raw, individual University of Colorado (CU) patient data are protected and not available due to data privacy laws. The processed data are available at reasonable request to the corresponding author. The Flatiron Health data that supported the findings of this study were originated by and are the property of Flatiron Health, Inc., which has restrictions prohibiting the authors from making the data set publicly available. Requests for data sharing by license or by permission for the specific purpose of replicating results in this manuscript can be submitted to.

## Abbreviations

AML: Acute Myeloid Leukemia
RWD: Real-world data
RWE: Real-world evidence
IPD: Individual-level patient data
PEM: Piecewise exponential model
PED: Piecewise exponential data
Cox-PH: Cox proportional hazard
PAM: Piecewise additive model
GAM: Generalized additive model
FAMES: **F**ederated **a**dditive **m**odel exploiting piecewise **e**xponential **s**urvival
DR: Demmler-Reinsch
CI: Confidence intervals
TPF: Truncated power function
TLS: Truncated linear splines
AIC: Akaike information criterion
BIC: Bayesian information criterion
WBC: White blood cell
ANC: Absolute neutrophil count
Hgb: Hemoglobin
Plt: Platelets
LDH: Lactate dehydrogenase
CKD: Chronic kidney disease
COPD: Chronic obstructive pulmonary disease
MDS: Myelodysplastic syndrome
VTE: Venous thromboembolism
rELN24: Refined European Leukemia Net, 2024
RRM: Refined risk model

## SUPPLEMENTAL MATERIALS

### Supplementary method

The following sections offer further details and numerical results relevant to the extension of the FAMES model. For the sake of notational brevity, we have refrained from introducing any new notations that share the same meaning as those presented in the main draft.

### Extension to multivariate smooth effects

The proposed FAMES model can be extended to an additive model with multidimensional smooth effects. Let ***U*** and ***V*** be two numerical covariates that are scaled via min-max standardization. For exposition, we assume bi-variate varying effects *f*(*u, v*) which is a flexible surface that captures effects of both additive and interaction effects. This smooth effect can be approximated using tensor product of truncated linear splines with each direction as

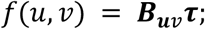

where ***B***_***uv***_ is a (*N* × *l*_*u*_*l*_*v*_) dimensional basis matrix and ***τ*** represents the stacked (*l*_*u*_*l*_*v*_ × 1) vector of unknown basis coefficients. Let ***B***_***i***,***uv***_ = ***B***_***i***,***u***_⨂ ***B***_***i***,***v***_ denote the Kronecker product of two basis functions where the elements of bases are arrayed in a wide column format for each subject. Using the similar intuition as above, the respective penalized likelihood function under the PED framework can be written as

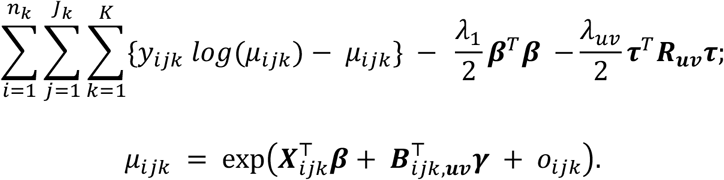

where all other terms bear the same meaning as before. Note we assume same smoothing parameter for controlling penalization in both *u* and ***v*** directions for ease of computation. If needed such assumption can be relaxed without loss of any generality; but this may increase computation time as optimization needs to be performed for each combination of parameters in the space. Here ***R***_***uv***_ corresponds to the Kronecker products between penalty matrices and is defined by ***R***_***uv***_ = (***D***_***u***_ ⨂ ***I***_***v***_) + (***I***_***u***_ ⨂ ***D***_***v***_). As before, both orthogonalization and diagonalization were accomplished through DR-transformation to ensure that the basis functions remain orthonormal and that the penalty terms associated with smooth effects are diagonal. Define the transformed basis functions and associated coefficients by 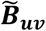 and ***θ***. The unknown parameters are estimated via the steps outlined in Table 1 and coefficients are transformed back into original format and two-dimensional varying coefficients are presented in a matrix format.

Subsequently, the variance-covariance matrix can be estimated as 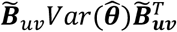 and subsequently, the corresponding surface of lower-and-upper bound of confidence intervals are generated. Note that the boundary regions of the bivariate surface may be susceptible to over-smoothing resulting in an artificially tight uncertainty bound due to limited local support or over-shrinkage. This underscores the need for careful interpretation of edge effects.

### Extension to multiple smooth effects

Without loss of generality, *f*(*u, v*) can be decomposed explicitly into additive (*f*_1_(*u*) and *f*_2_(*v*)) and interaction (*f*_12_(*uv*)) effects where each term is individually approximated using basis function expansion, and the DR-transformation is applied separately to each of the three terms. Subsequently, estimation is carried out via the penalized approach as previously described. Furthermore, the model can be extended to incorporate smooth interaction effects involving more than two dimensions and can be adapted for models where a dimension corresponds to a categorical feature. Using the similar intuition as above, the model can be extended to adjust for multiple smooth effects 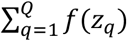 simultaneously; details of such modeling steps are stipulated in the Supplementary Table 1. The resulting penalized likelihood function with multiple smooth effects can be expressed as below-

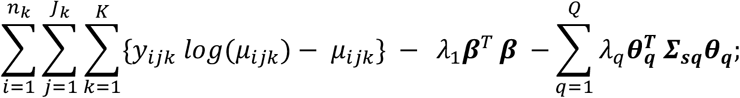

where, *λ*_*q*_′*s* correspond to tuning parameter controlling the goodness of fit and smoothness of curves. To reduce the computational burden, we assume, though restrictive in nature, same smoothing parameters for all smooth effects such as 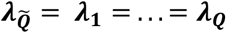.

---

**Supplemental Figure 1.**
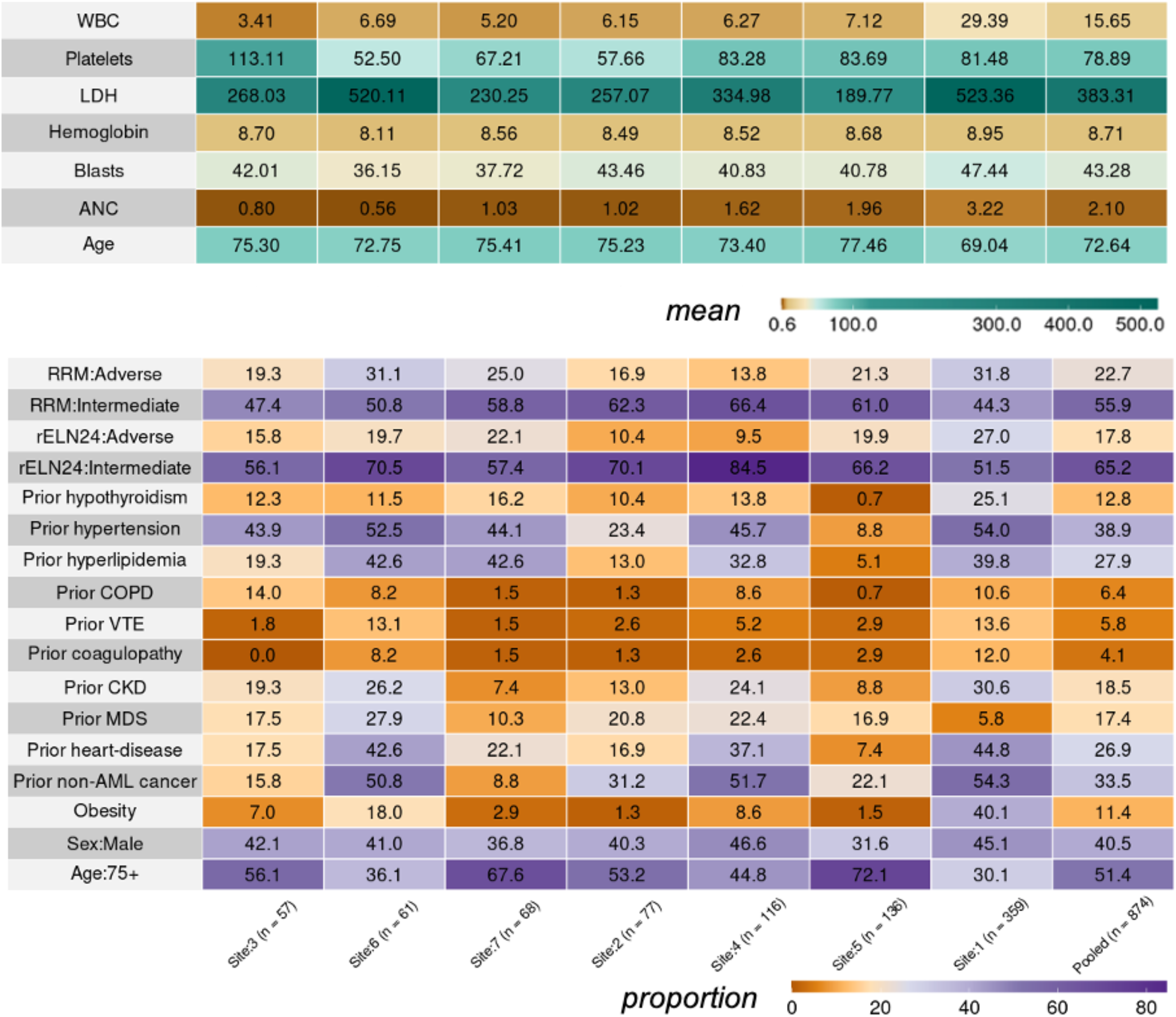
Summary statistics of features across 7 sites. Mean values are reported for continuous variables and prevalence rates are provided for categorical features.

**Supplementary Figure 2.**
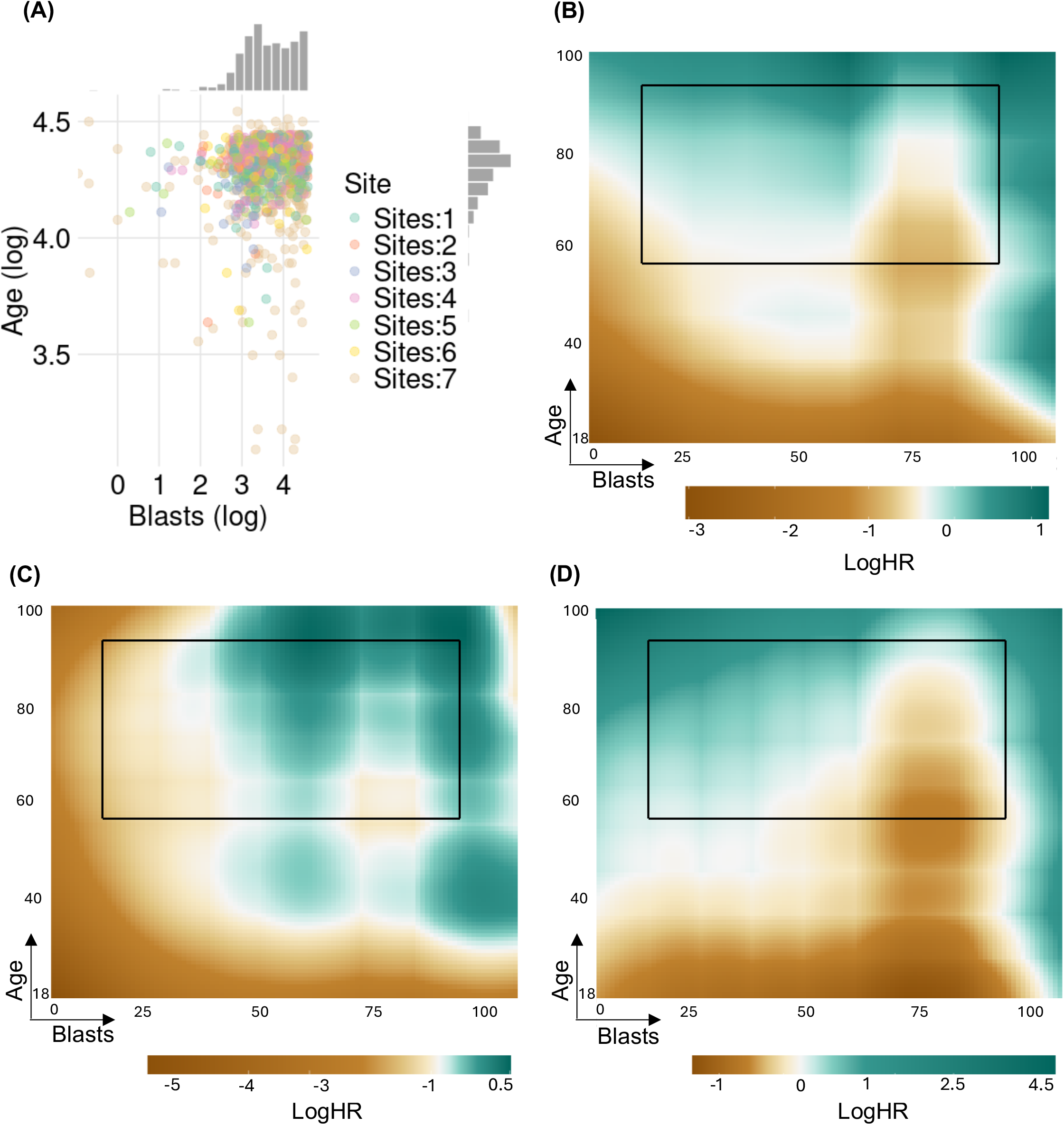
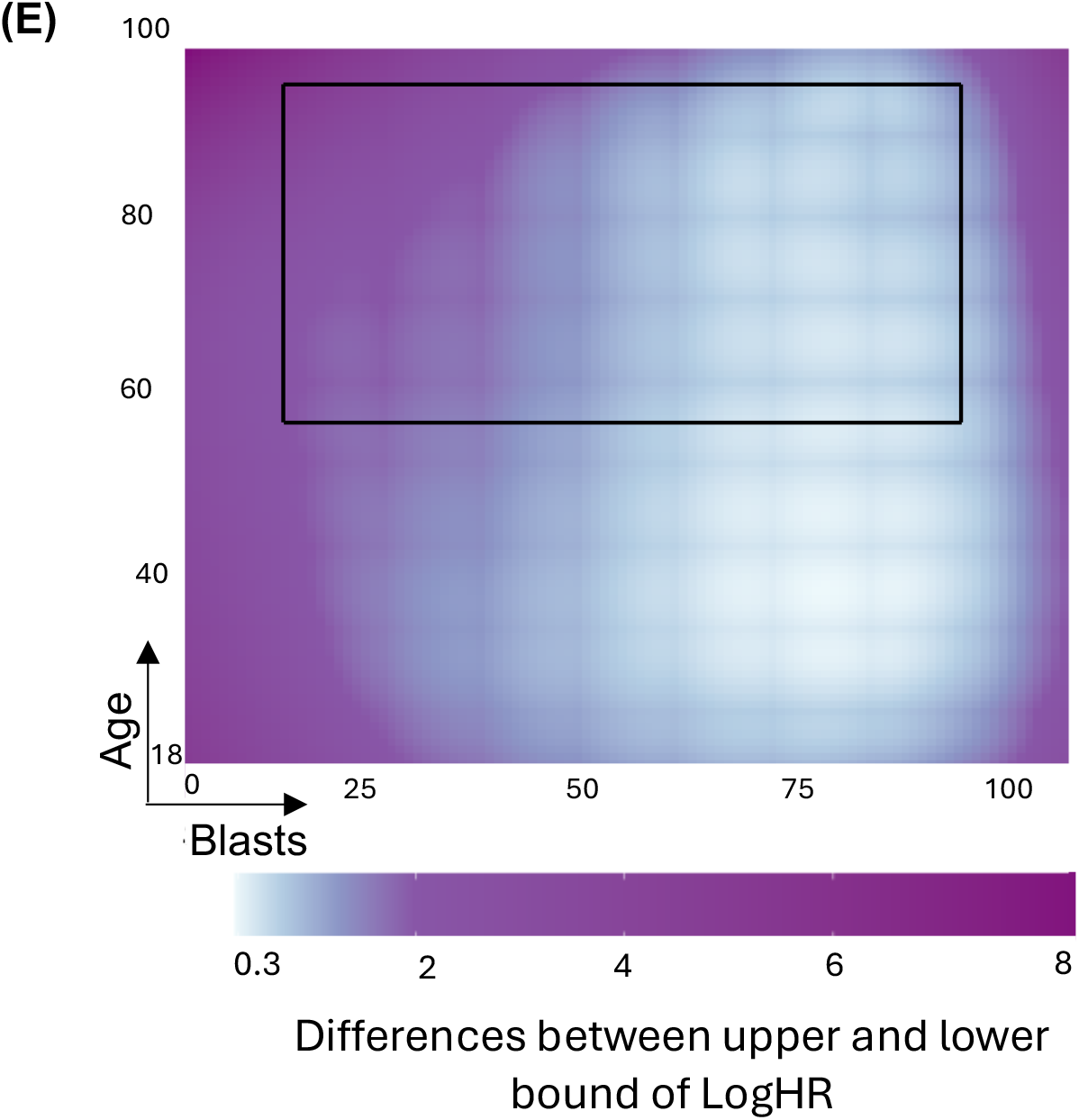
Estimates of bivariate smooth effects. Reported are the estimates of interaction effects (log hazard ratio) between age and blasts that are uniformly distributed within the minimum and maximum values of the corresponding features. Heatmaps correspond to (A) distribution of age and blasts values across 7 sites, (B) point-estimates, (C) lower-bound and (D) upper-bound of 95% CI, and (E) width of confidence intervals. The highlighted box corresponds to the area with high concentration of data for age = {60, 85} and blasts = {20, 90}.

**Supplementary Table 1.**
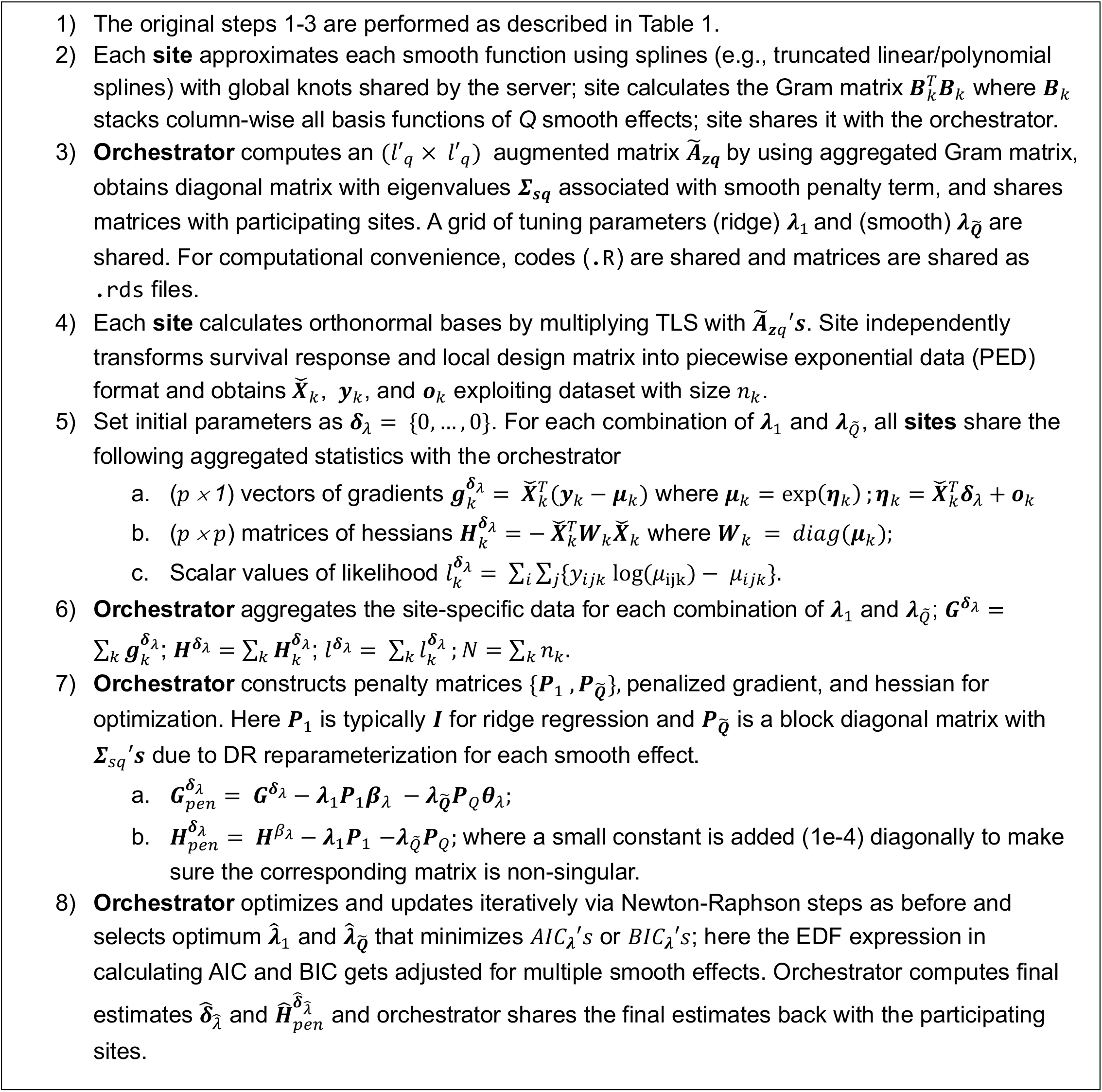
An overview of estimation process in FAMES with multiple smooth effects.

## References

1 Chai, H. et al. A decentralized federated learning-based cancer survival prediction method with privacy protection. Heliyon 10, e31873 (2024). 10.1016/j.heliyon.2024.e31873

2 Cai, Z. et al. Federated Deep Learning Enables Cancer Subtyping by Proteomics. Cancer Discovery 15, 1803–1818 (2025). 10.1158/2159-8290.Cd-24-1488

3 Han, L., Hou, J., Cho, K., Duan, R. & Cai, T. Federated Adaptive Causal Estimation (FACE) of Target Treatment EYects. J Am Stat Assoc (2025). 10.1080/01621459.2025.2453249

4 Ogier du Terrail, J. et al. FedECA: federated external control arms for causal inference with time-to-event data in distributed settings. Nature Communications 16, 7496 (2025). 10.1038/s41467-025-62525-z

5 Banerjee, S. et al. dsSurvival: Privacy preserving survival models for federated individual patient meta-analysis in DataSHIELD. BMC Research Notes 15, 197 (2022). 10.1186/s13104-022-06085-1

6 Imakura, A., Tsunoda, R., Kagawa, R., Yamagata, K. & Sakurai, T. DC-COX: Data collaboration Cox proportional hazards model for privacy-preserving survival analysis on multiple parties. Journal of Biomedical Informatics 137, 104264 (2023). 10.1016/j.jbi.2022.104264

7 Rollo, C. et al. SYNDSURV: A simple framework for survival analysis with data distributed across multiple institutions. Computers in Biology and Medicine 172, 108288 (2024). 10.1016/j.compbiomed.2024.108288

8 Miao, G. et al. Learning from vertically distributed data across multiple sites: An eYicient privacy-preserving algorithm for Cox proportional hazards model with variable selection. Journal of Biomedical Informatics 149, 104581 (2024). 10.1016/j.jbi.2023.104581

9 Westers, F., Leder, S. & Tealdi, L. Horizontal federated learning and assessment of Cox models. Frontiers in Digital Health Volume 7 - 2025 (2025). 10.3389/fdgth.2025.1603630

10 Islam, N. et al. Federated penalized piecewise exponential model for horizontally distributed survival data: FedPPEM. medRxiv, 2026.2002.2011.26346054 (2026). 10.64898/2026.02.11.26346054

11 Luo, C. et al. ODACH: a one-shot distributed algorithm for Cox model with heterogeneous multi-center data. Scientific Reports 12, 6627 (2022). 10.1038/s41598-022-09069-0

12 Andreux, M., Manoel, A., Menuet, R., Saillard, C. & Simpson, C. Federated Survival Analysis with Discrete-Time Cox Models. ArXiv abs/2006.08997 (2020).

13 Duan, R. et al. Learning from local to global: An eYicient distributed algorithm for modeling time-to-event data. J Am Med Inform Assoc 27, 1028–1036 (2020). 10.1093/jamia/ocaa044

14 Wood, S. Generalized Additive Models. Annual Review of Statistics and Its Application 12 (2024). 10.1146/annurev-statistics-112723-034249

15 Swenne, A., Intemann, T., Moreno, L. A. & Pigeot, I. Federated generalized additive models for location, scale and shape. BMC Medical Research Methodology 25, 276 (2025). 10.1186/s12874-025-02735-7

16 Hu, F. et al. dGAMLSS: an exact, distributed algorithm to fit Generalized Additive Models for Location, Scale, and Shape for privacy-preserving population reference charts. Bioinformatics 42 (2026). 10.1093/bioinformatics/btaf625

17 Islam, N. et al. Development of a dynamic counterfactual risk stratification strategy for newly diagnosed acute myeloid leukemia patients treated with venetoclax and azacitidine. medRxiv, 2024.2011.2025.24317750 (2024). 10.1101/2024.11.25.24317750

18 Islam, N. et al. A simplified risk model for pretreatment stratification of newly diagnosed acute myeloid leukemia patients treated with venetoclax and azacitidine. medRxiv, 2024.2012.2002.24318344 (2024). 10.1101/2024.12.02.24318344

19 Lachowiez, C. A. et al. Refined ELN 2024 risk stratification improves survival prognostication following venetoclax-based therapy in AML. Blood 144, 2788–2792 (2024). 10.1182/blood.2024026925

20 Bender, A., Groll, A. & Scheipl, F. A generalized additive model approach to time-to-event analysis. Statistical Modelling 18, 299–321 (2018). 10.1177/1471082x17748083

21 Demarqui, F., Loschi, R., Dey, D. & Colosimo, E. A class of dynamic piecewise exponential models with random time grid. Journal of Statistical Planning and Inference 142, 728–742 (2012). 10.1016/j.jspi.2011.09.006

22 James, G., Witten, D., Hastie, T. & Tibshirani, R. An Introduction to Statistical Learning: with Applications in R. (Springer Publishing Company, Incorporated, 2014).

23 Williams, C. & Shawe-taylor, J. The stability of kernel principal components analysis and its relation to the process eigenspectrum. Advances in neural information processing systems 15 (2002).

24 Demmler, A. & Reinsch, C. Oscillation matrices with spline smoothing. Numerische Mathematik 24, 375–382 (1975).

25 Tibshirani, R. & Wasserman, L. Nonparametric regression. Statistical Machine Learning, Spring (2013).

26 Wood, S. N. mgcv: GAMs and generalized ridge regression for R. R news 1, 20–25 (2001).

